# SIDIR: Extending SIR with Detected and Isolated Populations for Pandemic Modeling

**DOI:** 10.1101/2020.07.20.20157834

**Authors:** Joe Garman, Sean MacAvaney, Eugene Yang, Ophir Frieder

## Abstract

We extend the Susceptible Infected Recovered (SIR) model to include Detected (D) and Isolated (I) compartments (SIDIR). SIDIR improves COVID-19 outbreak behavior modeling by identifying infected populations as subsequently quarantined to reduce the spread of the infection (either in a hospital or self-quarantined). We present the model and provide a case study on COVID-19. The model estimates undetected cases (i.e., those infected but unconfirmed) and extrapolates when no additional undetected active (rogue) cases will remain, potentially guiding policy decisions that help control the spread of COVID-19 and future epidemics. A live demonstration of SIDIR on COVID-19 is available at: http://ir.cs.georgetown.edu/sidir.

## 1 Introduction

Paramount to combating infectious diseases is the accurate forecasting of their progression. Towards that end, since the early 1900s, epidemiological compartmental models that simplify the mathematical modeling of disease spread and duration were developed. Scenarios can be introduced and their effects studied. A simple and popular model among them is the Susceptible, Infectious, Recovered (SIR) model [1], and many additional expanded/modified models based on SIR exist. These expanded/modified models incorporate additional compartments such as, but not limited to, exposure, maternally inherited immune antibodies, death, and non-suffering carriers. Compartments are linked with progression paths, each compartment representing a potential state of population members. Individuals progress from compartment to compartment representing disease flow.

We introduce a subsuming variation of the SIR model called Susceptible, Infected, Detected, Isolated, Resolved (SIDIR). SIDIR deploys logically four primary compartments, namely: “Susceptible” - individuals that may contract the disease; “Infected” - individuals that are infected with the disease independent of exhibiting symptoms; “Isolated” - individuals that, regardless of their disease severity, are isolated at home or in treatment care centers; and “Resolved” - individuals that recovered or passed. We evaluate SIDIR on COVID-19, and demonstrate that it identifies the extent of case under-counting and estimates when undetected active cases will no longer exist in the population. A live demonstration of SIDIR on COVID-19 is available.^1^

## 2 Related Work

Epidemiologists have long relied on mathematical models to predict various epidemic characteristics, such as the infection and mortality rates. Compartmental models accomplish such predictions by dividing the population into sub-groups and extrapolating behavior from collected data. A foundational compartmental model is SIR that divides the population into Susceptible, Infectious, and Recovered compartments [1]. Though popular, SIR, given its simplicity, fails to accurately model disease specifics. Thus, numerous variations were proposed, including the introduction of an Exposed (E) compartment (SEIR). For comprehensive model reviews, we refer the reader to [2, 3, 4].

A model similar to our SIDIR is the SIQRD model, which introduces Quarantined and Deceased compartments [5]. Although conceptually similar, SIDIR neither requires nor assumes that the infected population can be measured directly and that when individuals are infected, they are promptly isolated (i.e., quarantined). As we show in Section 4, this assumption relaxation more closely matches the response pattern of current respiratory illnesses, such as COVID-19.

Numerous others have and are investigating mathematical models during the COVID-19 pandemic [6, 7, 8, 9, 10, 11, 12, 13]. The SuEIR model includes undetected diagnoses [6]. However, SuEIR assumes that detected and undetected cases have the same infection rate. Our model differs by treating detected cases as isolated, with a limited infection rate. Li [7] uses a SEIR model parameterized based on state responses and social contact rates. Rather than relying on these characteristics (which can vary from state-to-state), SIDIR predicts transmission rates based on confirmed cases and deaths. Another variant of the SIR model proposed adapts the rates over time [8]. Gu [9] uses a SEIR model with a grid search approach to find appropriate rates. Sheldon and Gibson [10] propose a Baysean SEIR model, with added compartments for Deaths and Hospitalized-and-will-die groups. In contrast, our model includes detected and isolated compartments. Others investigated the incorporation of mobility information (e.g., travel, commutes) into the SEIR model [11] or a stochastic model [12] for modeling the COVID-19 pandemic. Murray [13] proposed a non-linear regression model based on social distancing measures and cell phone usage. Our model is able to provide forecasts without utilizing this additional information. Unlike others that focus on predicting the total number of COVID-19 cases (e.g., [14, 15]), the SIDIR model specifically focuses on identifying undetected cases based on the difference between actual deaths and the known infection death rate in particular regions. Based on this, SIDIR is able to predict when there will be no remaining undetected cases.

## 3 SIDIR Model

Susceptible, Infected, Detected, Isolated, Resolved (SIDIR) subsumes the SIR model. As shown in Figure 1, SIDIR deploys logically four primary compartments with transitions between them. The “Susceptible”(*S*) compartment, in blue, comprises of individuals that may contract the disease. In addition to the local resident “susceptible” population, there is the possibility of imported cases, i.e., infected individuals moving into an area. These “imports” are required to initiate spread, play a meaningful role when the number of infected are low, e.g., starting and ending phases of a pandemic, but are relatively low in terms of count and significance during the primary stages of the pandemic.

**Figure 1:**
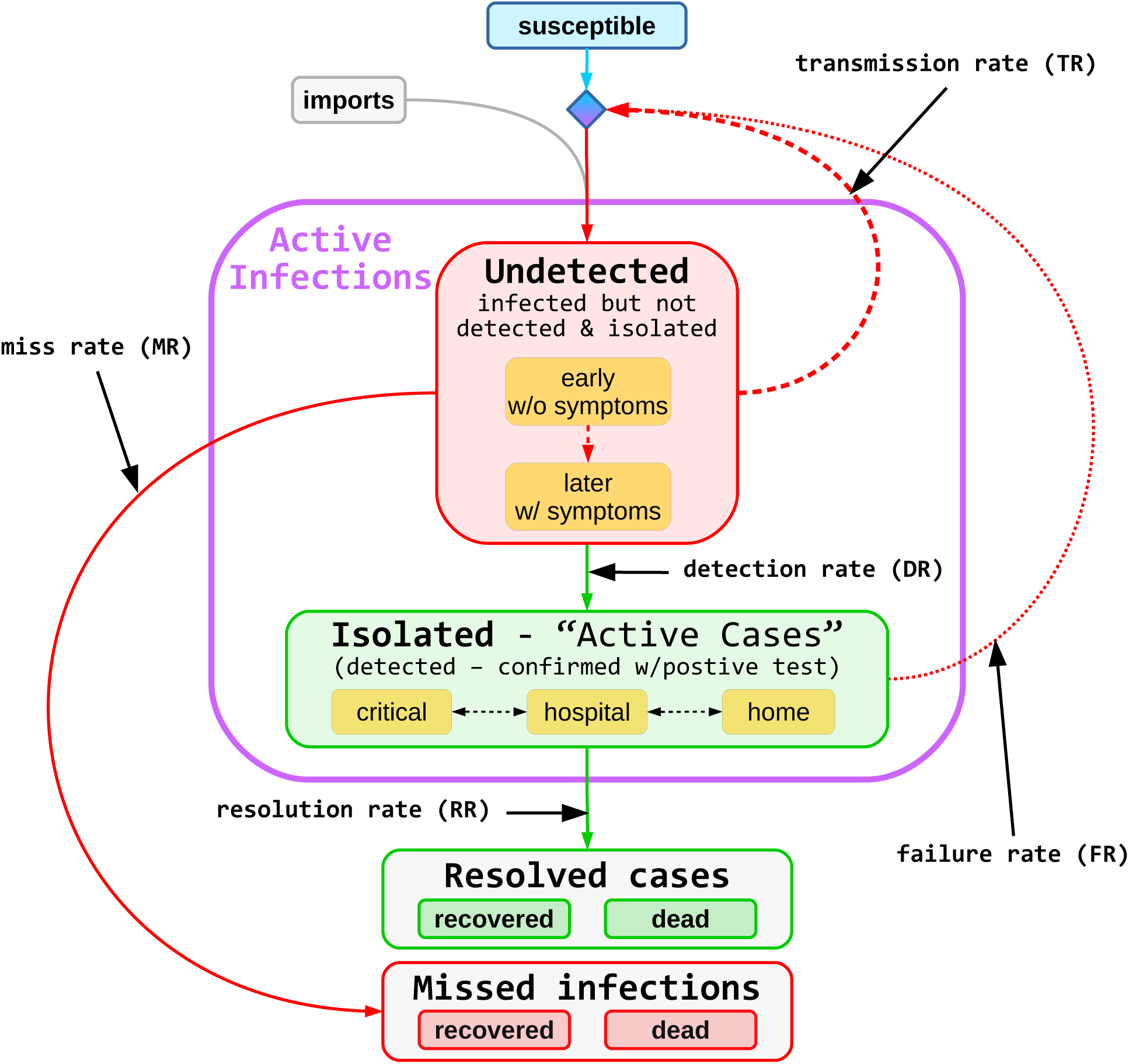
Model diagram. Green represents data: active cases, recoveries, and deaths. Red represents hidden values: undetected and missed infections. Undetected individuals are not isolated; so they are more contagious than isolated individuals. Missed infections are not reported as infections. That is, they account for the under counting of the reported infected or dead. SIDIR assumes that community transmissions dominate over imported cases, but allows any number of imports. Transmission can only occur if there is sufficient susceptible population.

Individuals infected with the disease, independent of exhibiting symptoms, are captured in the “Undetected”(*U*) compartment shown in red. As seen, these individuals initially are merely carriers of the infection and are symptom free; later as the infection progresses, they may exhibit associated effects. Regardless, none of the individuals within the “Undetected” compartment are detected or verified as having the infection.

Multiple transitions from the “Undetected” compartment exist. Individuals within this compartment infect others, with some “transmission rate” (TR, *β*), and remain within the compartment. They transition from the “Undetected” compartment by being detected as having the disease, with some “detection rate” (DR, *κ*), or resolving their situation either positively by recovering or negatively by succumbing to the disease. If and when detected, with a given “detection rate” (DR), they transition to the “Isolated”(*D*) compartment indicated in green. Otherwise they transition to the “Missed Infections”(*M*) compartment, which could be either recovered or dead, indicated in red by a “miss rate”(MR, *ω*). The unaccounted, namely those that transition via the miss rate transition, result in the under-representation of the disease impact: infection rate and mortality rate.

The “Isolated”(*D*) compartment consists of all individuals verified, via testing, as infected regardless of their disease severity. They are all in isolation either at home or in treatment care centers. Their condition can vary over time, and thus, they might migrate between home and treatment care centers, repetitively. Regardless of their physical location, they remain in the “Isolated” compartment until either they recover or succumb and transition to the “Resolved” compartment, with the rate of “resolution rate”(RR, *γ*).

While in the “Isolated”(*D*) compartment, some of these individuals inadvertently infect others. Such disease spread represents the infection of family members while isolated at home and care-givers while in treatment centers. This infection process is represented by the “failure rate” (FR, *η*), which accounts for transmission of the infection from “Isolated” (D) individuals to the “Susceptible” individuals resulting in additional “Undetected” (U) infections. The “failure rate” is typically much lower than the “transmission rate”.

Using SIR-style notation, we define the force of infection *λ* as the per capita rate of the susceptible individuals being infected. Since the population having the ability to transmit (*N*) determines force, assuming isolation is successful, transmission occurs only from the undetected individuals, and *lambda* is the function of the “undetected” compartment(*U*). Therefore,

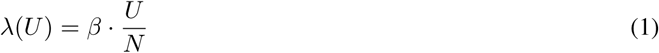

where *β* is the rate of transmission as defined above.

The dynamic of the model is defined as follows.

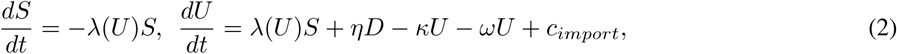

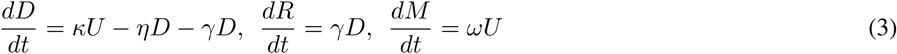

where *c*_*import*_ is the time-independent constant of the import rate.

Given the current pandemic, we focus on the situation where most of the population is still susceptible, targeting the spread of the virus at this point. Therefore, we assume *S/N ≈* 1 throughout. Consequently, *λ*(*U*) can be rewritten as *βU/S* and the dynamic of *S*, and *U* can be rewritten as:

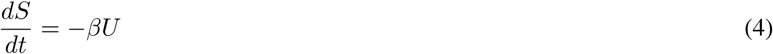

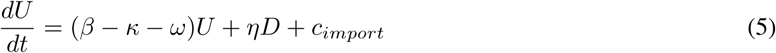

For modeling purposes, we can rewrite the above formulation in an iterative form. At time *t*, the changes of the compartments follow,

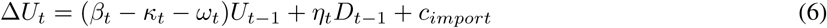

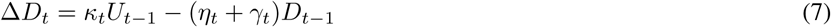

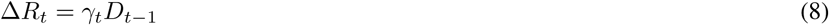

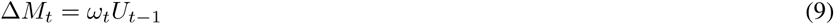

where each of the rates can be time-dependent, as measurements change over time. Therefore, let *I* be the size of infected population, the change in *I* is

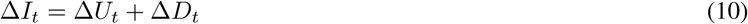

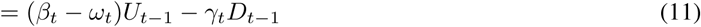

As some of the variables can be observed, the rates and other latent variables can be estimated. In the following sections, we discuss value estimation.

### 3.1 Limitations and Simplifying Assumptions

We acknowledge that SIDIR makes simplifying assumptions that potentially limit the model. First, the model fails to consider cases in which resolved cases re-infect the population. Such infection can occur either from recovered or deceased individuals; we assume such infections as infrequent. The model also ignores recovered individuals becoming susceptible again (and thus could again become infected). For COVID-19, there is still insufficient evidence about how long the virus antibodies last [16, 17]. For simplicity, the model assumes that the TR from undetected cases will dominate the effects of either of these situations.

## 4 Case Study: COVID-19 Pandemic

To validate the SIDIR model, we evaluate the model’s ability to forecast the current COVID-19 pandemic. Specifically, we demonstrate how it can be used to predict undetected cases (which we can extrapolate from case fatality rates in a given region), and ultimately the date at which there will be no more undetected active cases (what we call Virus Victory Day, VVD).

### 4.1 Observed Parameters

We explore the SIDIR model based on the observed COVID-19 active cases, death counts, and Case Fatality Rate (CFR) for a given region, the typical timeline of the virus and its average Infection Fatality Rate (IFR).

#### 4.1.1 Active Cases and Deaths

We utilize data curated by the Johns Hopkins University Center for Systems Science and Engineering (JHU CSSE), made available via their interactive dashboard [18].^2^ The dataset consists of total counts for confirmed cases, deaths, recovered cases, and remaining active cases at a daily granularity. We use this data source for active case and death counts, disregarding the other values. The values are partitioned by region—such as country, state, provinces, etc. In the United States, values by individual city or counties are also available. The dataset reflects the latest available data from a variety of governmental and intergovernmental organizations, including but not limited to: the World Health Organization (WHO), the mainland China CDC, the Taiwan CDC, the United States CDC, the European Centre for Disease Prevention and Control (ECDC). Data are extended and corroborated with other sources, such as DXY,^3^ BNO News,^4^ and the COVID Tracking Project.^5^

#### 4.1.2 Disease Timeline

COVID-19 progresses through a typical infectious disease timeline (overview in Table 1). After individuals are exposed to the disease, they may become infected, and later become infectious (i.e., can infect others). They then may show symptoms, which could lead to detection via a test. A positive test will lead to either self-isolation or isolation in a hospital. Those with mild cases can recover quickly (mean of 21 days in our model). Those with severe cases take longer to recover or result in death.

**Table 1:** Timeline of COVID-19 infection.

| Event | Means from literature | Mean for our model |
| --- | --- | --- |
| Exposure | - | - |
| Infection | - | 0 |
| Infectious | 0–2 | - |
| Symptomatic | 5–7 | - |
| Detection | 9–15 | 12 |
| Hospitalization | 12–16 | - |
| Recovery (mild cases) | 21 | 21 |
| Death | 21–25 | 23 |
| Recovery (severe cases) | 28 | - |

We simplify this timeline by considering three ranges: the time between infection and detection, the time between infection and recovery, and the time between infection and death. To further simplify, we only consider recovery of mild cases. We represent the ranges for the exposure-to-detection, exposure-to-recovery, and exposure-to-death as scaled gamma distributions

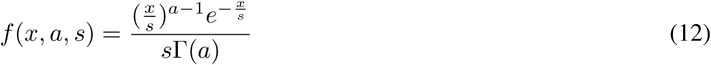

parameterized based on typical values found in literature [19, 20, 21]. The parameters are given in Table 2, and the probability distribution functions of each are plotted in Figure 2.

**Table 2:** Parameters of exposure-to-detection, exposure-to-recovery, and exposure-to-death gamma distributions.

| Distribution | $a$ (mean/shape) | $s$ (shape) |
| --- | --- | --- |
| exposure-to-detection | $\frac{12}{2} = 6.0$ | 2 |
| exposure-to-recovery | $\frac{21}{2} = 10.5$ | 2 |
| exposure-to-death | $\frac{23}{7} = 3.3$ | 7 |

**Figure 2:**
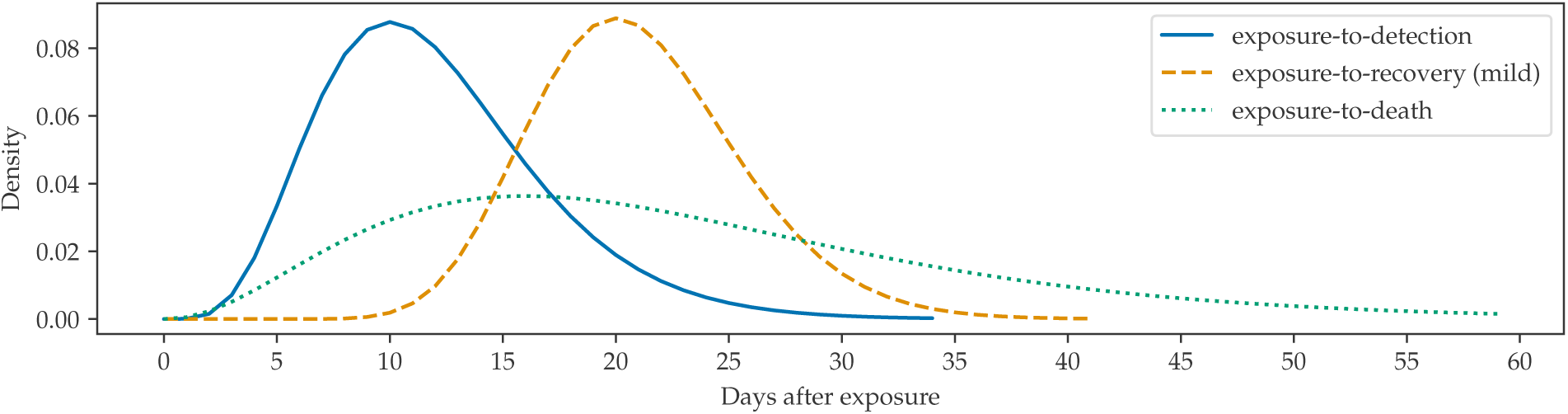
Probability distribution functions of exposure-to-detection (solid blue), exposure-to-recovery (dashed orange), and exposure-to-death (dotted green). The parameters of the distributions (Table 2) were selected based on values found in the literature (Table 1).

#### 4.1.3 Infection Fatality Rate (IFR) and Case Fatality Rate (CFR)

To estimate rates of the model (e.g., TR, DR, and RR), we use the observed Infection Fatality Rate (IFR). Note that this differs from the Case Fatality Rate (CFR) in that it also includes the infections that were not detected. CFR depends on region (due to, e.g., quality of healthcare and prevalence of testing) and can be easily calculated based on case and death counts for a particular region (after accounting for the lag between detection and death). Determining the IFR from CFR requires several corrections, including case under-counting due to insufficient testing, adjustments by age, comorbidities (i.e., multiple pre-existing conditions), differences in medical care, and under-counting of deaths. From prior literature [20, 22, 23, 24, 25, 26, 27], we use an IFR of 1.2%.

### 4.2 Estimating the Extent of Case Undercounting

Armed with confirmed COVID-19 case counts by date, the infection-to-detection distribution, we can estimate the date of *infection* for the confirmed COVID-19 cases in a particular region. To do this, we first smooth the data for a particular region using two iterations of the Savitzky-Golay filter. We present this for Washington, DC, in Figure 3. We observe that when projecting the estimated infection date to cases and deaths, the predicated counts align with observed case counts.

**Figure 3:**
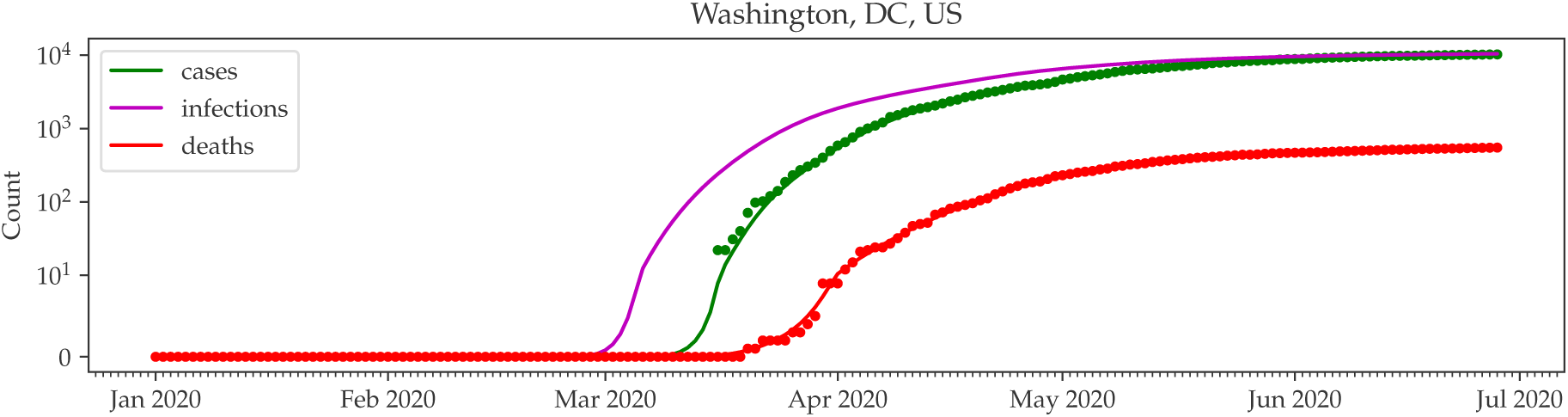
Cumulative case counts, death counts, and estimated date of infections for Washington, DC (log scale). Smoothed case and death counts are indicated with solid lines. Case counts based on the estimated infection counts are marked with a magenta x.

If all cases were properly detected, the region’s CFR would be similar to the virus’ global IFR. However, this is often not the case, implying a substantial case undercount. To estimate the extent of this undercount, we first apply the IFR to the predicted infection counts. The predicted deaths given the IFR for Washington, DC, and New Zealand are shown in Figure 4. For Washington, DC, the predicted deaths fall substantially below the observed COVID-19 death counts, indicating an undercount of cases. In New Zealand, where more extensive testing has taken place, the predicted deaths more closely mirror the actual death counts.

**Figure 4:**
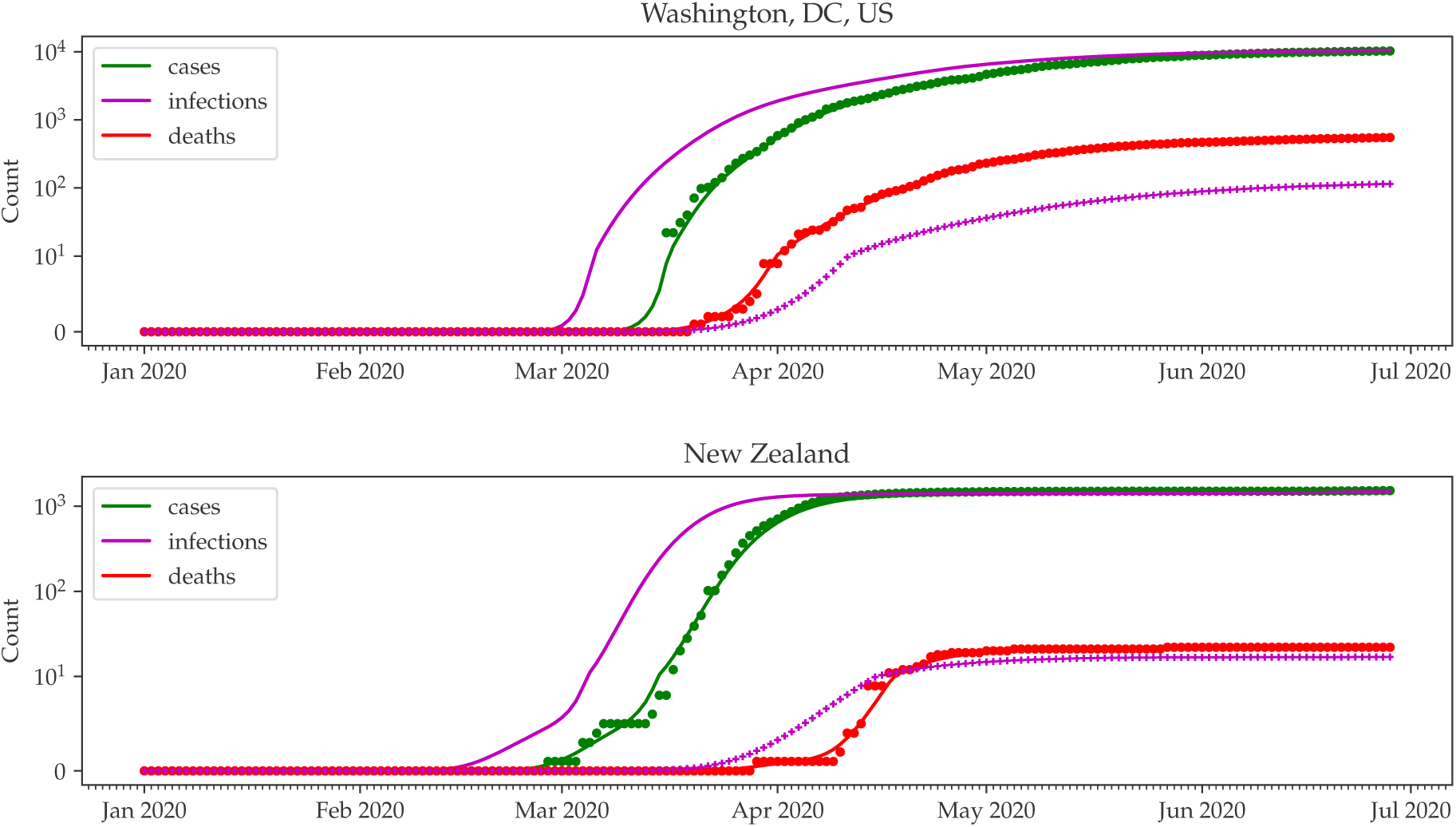
Predicted cumulative death counts for (a) Washington, DC, and (b) New Zealand (magenta x). Note that since this is substantially lower than the observed death counts in Washington, DC, case undercounting has likely occurred. In contrast, in New Zealand, the predicated deaths more closely mirror the observed deaths.

To quantify the extent to which cases were undercounted, the infection-to-death distribution can be applied to the IFR-predicted death counts. In the case of Washington, DC, we observe that the majority of cases are undetected (Figure 5). The number of missed cases is the difference between the total cases and the observed total active cases.

**Figure 5:**
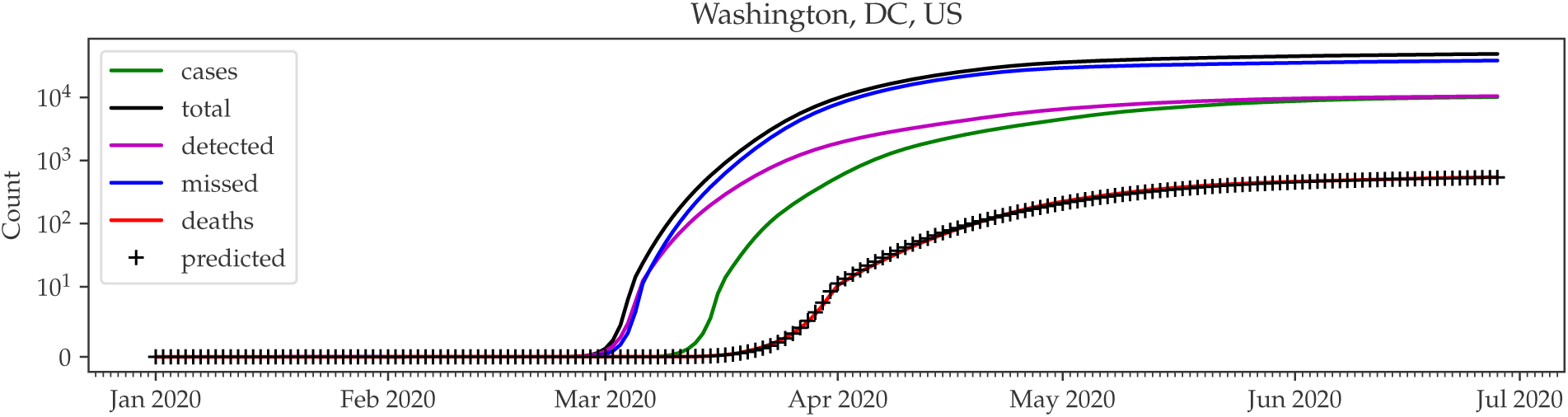
Predicted total cases in Washington, DC.

As a sanity check, we use the predicted total cases and IFR to predict the deaths by date (black pluses in Figure 5). We find that these predictions align well with the actual observed death counts.

### 4.3 Estimating Active Undetected Infections

Through the total predicted cases, detected cases, and deaths, we can model the changing TR, DR, and MR over time. See Figure 6 for the rates in Washington, DC. Note that we clip TR to 0.5. We observe that outbreaks typically follow a pattern of high transmission at the start due to superspreaders, then decline in TR due to the law of small numbers. Policies, such as social distancing and quarantining, can further reduce the TR, and the easing of these policies potentially increase TR. DR is a function of testing. In the case of COVID-19 in Washington, DC, the testing is clearly insufficient to identify all cases because it remains much lower than the MR.

**Figure 6:**
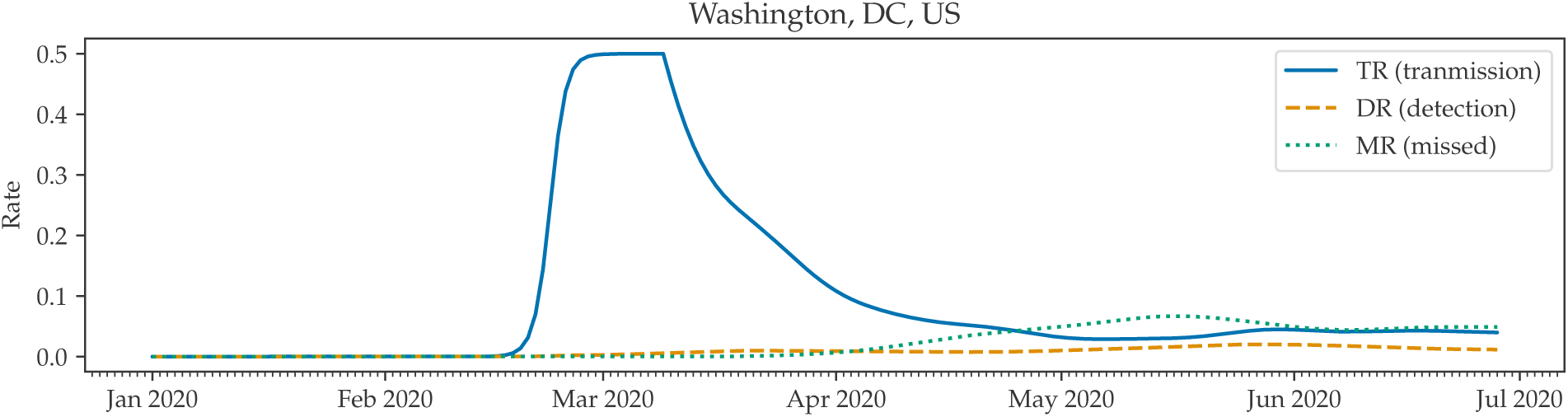
Transmission Rate (TR), Detection Rate (DR), and Miss Rate (MR) for COVID-19 in Washington, DC.

Given the aforementioned rates, the remaining number of active but undetected “rogue” cases (i.e., possibly asymptomatic cases for which the patient has not yet gained immunity) can be derived. Rogue cases are problematic because the individuals may be spreading the infection without knowing it, leading to a higher TR. We plot this value for Washington, DC, in Figure 7. Note that as individual immunity is gained, rogue cases can decline over time with a sufficiently low TR.

**Figure 7:**
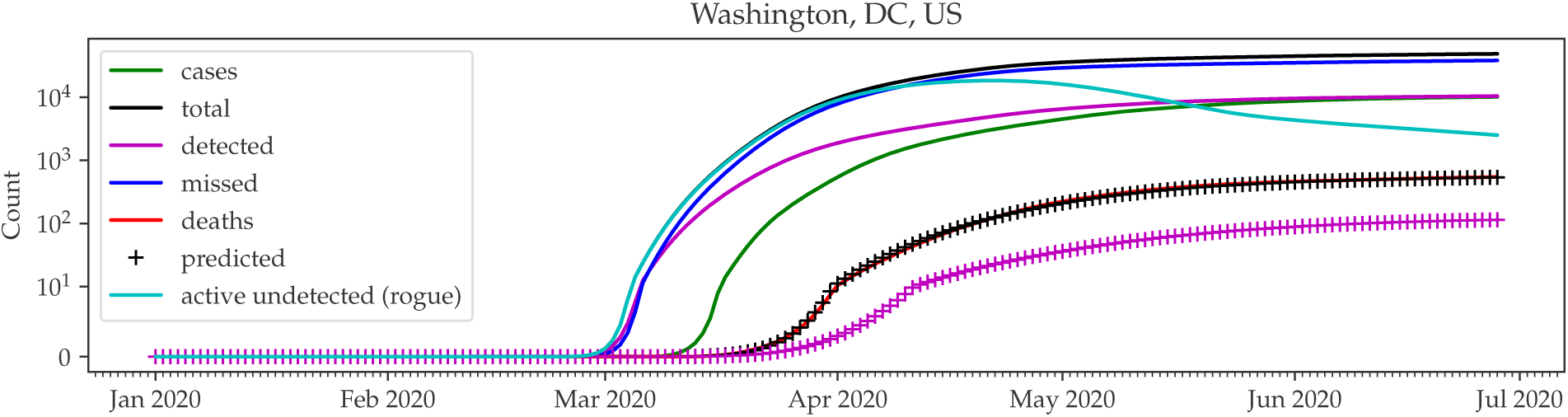
Rogue cases in Washington, DC. Note that the rogue cases are beginning to fall.

### 4.4 Projecting the End of Pandemic

We now use the predicted rogue cases to estimate the end of the pandemic. To do this, we simply extrapolate the exponential trend of the rogue cases in the past week. When this value reaches 0, there will be no rogue cases remaining, and thus all cases are either active (and thus isolated), or resolved. In this setting, the only new infections can come as a result of the Failure Rate or additional imports, which can be more easily managed. We call the specific date at which there are no remaining rogue cases Virus Victory Day (VVD). We present the VVDs as of June 28, 2020, for several regions in Table 3. Indeed closely matching the prediction shown, as of July 16, 2020, Taiwan detected only 2 positive cases since its VVD on July 4, 2020.

**Table 3:** Virus Victory Dates (VVDs) for several regions, as of June 28, 2020. Regions marked with N/A have a positive trajectory of rogue cases.

| Region | VVD |
| --- | --- |
| Taiwan | 4 Jul 2020 |
| Washington, DC, US | 16 Jul 2021 |
| Chicago (Cook County), IL, US | 8 Oct 2021 |
| New York City, NY, US | 24 May 2029 |
| Orlando (Orange County), FL, US | N/A |

## 5 Discussion and Conclusion

We extended the SIR model to incorporate Detected (D) and Isolated (I) compartments (SIDIR). We demonstrated SIDIR modeling capability for pandemics because it represents realistic responses, i.e., that detected individuals are isolated from the remainder of the population. By allowing for infection, detection, and death rates to vary over time, SIDIR can evaluate effectiveness of certain policies, such as social distancing and increased testing. Furthermore, it allows the computation of the end-of-epidemic (Virus Freedom Day), based on when there are no remaining undetected active cases.

We fully understand that the introduction of new shelter-in-place or travel restrictions or contract-tracing tools (e.g., [28]) can alter expected and observed behavior or detection. That said, we hope that this tool can be beneficial for evaluating the effect of policies regarding epidemics such as COVID-19.

## Data Availability

All data is publicly available from the COVID-19 Data Repository by the Center for Systems Science and Engineering (CSSE) at Johns Hopkins University.

https://github.com/CSSEGISandData/COVID-19

## Footnotes

1 http://ir.cs.georgetown.edu/sidir

2 JHU CSSE aggregate data available at https://github.com/CSSEGISandData/COVID-19

3 http://3g.dxy.cn/newh5/view/pneumonia

4 https://bnonews.com/index.php/2020/02/the-latest-coronavirus-cases/

5 https://covidtracking.com/about-project

## Notes

### Competing Interest Statement

The authors have declared no competing interest.

### Funding Statement

This research received no external funding.

### Author Declarations

Georgetown IRB. Since all the data to be gathered for the study is publicly available, the study does not meet the definition of research on human subjects, and does not need to be submitted for IRB approval.

